# Examining the association between the *FTO* gene and neuroticism reveals indirect effects on subjective well-being and problematic alcohol use

**DOI:** 10.1101/2024.02.20.24303076

**Authors:** Wenjie Cai, Yvonne Forsell, Catharina Lavebratt, Philippe A. Melas

## Abstract

Associations between the fat mass and obesity-associated (*FTO*) gene and obesity are well-established. However, recent studies have linked *FTO* to addiction phenotypes and dopaminergic signaling, thus suggesting broader psychiatric implications. We explored this assumption by conducting a phenome-wide association study (PheWAS) across 4,756 genome-wide association studies (GWASs), identifying 26 psychiatric traits associated with *FTO* at the multiple-corrected significance level. These traits clustered into four categories: substance use, chronotype/sleep, well-being, and neuroticism. To validate these findings, we analyzed a functionally suggestive *FTO* variant (rs1421085) in a separate cohort, examining its impact on (i) alcohol use based on the Alcohol Use Disorders Identification Test (AUDIT), (ii) subjective well-being based on the WHO (Ten) Well-Being Index, and (iii) neuroticism based on Schafer’s Five Factor Model (FFM) or the Karolinska Scales of Personality (KSP). Our results confirmed a direct association between rs1421085 and neuroticism that was independent of age, sex, alcohol use, body mass index (BMI), and childhood adversities. Interestingly, while no direct association with alcohol intake was observed, both cross-sectional and lagged longitudinal mediation analyses uncovered indirect relationships between rs1421085 and problematic alcohol use (AUDIT-P), with increased neuroticism acting as the intermediary. Mediation analyses also supported an indirect effect of rs1421085 on lower well-being through the pathways of increased neuroticism and BMI. Our study is the first to validate a direct association between *FTO* and neuroticism. However, additional studies are warranted to affirm the causal pathways linking *FTO* to well-being and alcohol use through neuroticism.

## Introduction

The gene encoding the alpha-ketoglutarate-dependent dioxygenase (*ALKBH9)*, popularly known as the fat mass and obesity-associated gene (*FTO*), is widely recognized for its role as a genetic factor predisposing to obesity ^1,2^. More recently, however, genome-wide association studies (GWASs) have reliably associated *FTO* with an elevated risk of various alcohol use traits, independent of body mass index (BMI) effects ^3-6^. Additionally, a GWAS examining multiple substance use disorders suggested that *FTO* may represent a broader risk factor for addiction ^7^. More specifically, single nucleotide polymorphisms (SNPs) within the first intronic region of *FTO* have been associated with alcohol use disorder (rs1421085) ^3^, alcohol consumption (rs62033408) ^3^, and problematic alcohol use (rs9937709, rs1421085) ^4,8^. Multitrait analysis of GWAS (MTAG) identified associations between *FTO* (rs7188250) and both maximum habitual alcohol intake and problematic alcohol use ^5^. The largest GWAS of drinking phenotypes so far, encompassing nearly 3.4 million individuals, found associations between *FTO* variants and weekly alcohol consumption (rs1421085, rs11642015, rs62048402, rs1558902) ^6^. Furthermore, a gene-based analysis within a multivariate GWAS assessing a general addiction risk factor, found that *FTO* was the most significant gene involved (fine-mapped SNP: rs1477196) ^7^.

While much of the *FTO* research has concentrated on obesity, its potential involvement in neuropsychiatric disorders is gaining traction, supported by both preclinical and translational studies ^9^. *FTO* encodes an m6A RNA demethylase, highly enriched in neurons, and plays a role in neurotransmission and memory formation, including adult neurogenesis ^10-16^. Notably, deactivation of the *Fto* gene in animal models disrupts D2 and D3 dopamine receptor-dependent control of neuronal activity and behavioral responses ^11^. Given the dopaminergic system’s extensive involvement in various psychiatric disorders ^17^, and the known comorbidity of substance use disorders with these conditions ^18^, we postulated that *FTO* may influence a broader range of psychiatric traits beyond the addiction phenotypes already identified. To explore this, we conducted a phenome-wide association study (PheWAS) and followed up with genotyping a functionally relevant SNP in a separate cohort. Our research not only proposes FTO as a potential target for psychiatric treatment but also emphasizes the importance of considering personality traits, such as neuroticism, in developing intervention strategies.

## Methods

### Phenome-Wide Association Study (PheWAS)

To investigate the broader implications of the *FTO* gene, we conducted a PheWAS utilizing the Atlas of GWAS Summary Statistics (GWAS Atlas, https://atlas.ctglab.nl/, last accessed November 9, 2023) ^19^. This analysis encompassed 4,756 GWAS datasets, covering 3,302 unique phenotypic traits. A PheWAS plot was generated to visualize associations for 791 traits meeting the nominal p-value cutoff of 0.05. To adjust for multiple comparisons, we applied a Bonferroni correction, setting the significance threshold at a p-value of 6.32e-5 (0.05/791).

### Replication cohort: cross-sectional and longitudinal data

Our replication phase leveraged data from PART, a longitudinal cohort study from Stockholm County, Sweden, which relies on self-administered questionnaires (validated by interviews with psychiatrists) to explore risk and protective factors for mental health ^20^. For cross-sectional analyses, and unless otherwise indicated, we extracted and analyzed data on demographics (age and sex), anthropometrics (BMI) and psychiatric-related traits (alcohol use, well-being, and neuroticism) from the third wave of PART (PART wave III; 2010–2011) ^21^, which was the only wave to comprehensively assess neuroticism using the Schafer’s Five Factor Model (FFM) personality scale (see below). For longitudinal mediation analyses, data on BMI, alcohol use, well-being, and a subset of neuroticism-related items from the Karolinska Scales of Personality (KSP; see below), were also extracted from the second wave of PART (PART wave II; 2001–2003) ^22^. Ethical approval for PART was obtained from the Karolinska Institutet’s review board (nr. 96-260 and 97-313 for questionnaire data, nr. 2004-528/3 for DNA data, and nr. 2009/880-31 for PART wave III), in line with the World Medical Association’s (WMA) Helsinki Declaration. All participants provided informed consent, and all described methods were carried out in accordance with relevant guidelines and regulations.

### Alcohol use measurement

Alcohol use was assessed in the PART study using the 10-item Alcohol Use Disorders Identification Test (AUDIT), which examines drinking habits and associated problems over the past 12 months, as previously described ^23,24^. The analysis included the full AUDIT scale (items 1–10), the AUDIT-C subscale focusing on consumption (items 1-3), and the AUDIT-P subscale targeting problematic usage (items 4–10).

### Subjective well-being measurement

Subjective well-being in the PART study was measured using the WHO (Ten) Well-Being Index, a derivative of the WHO (Bradley) Subjective Well-Being Inventory Index, as previously described ^25-27^. This 10-item scale ranges from 0 (never) to 3 (always), reflecting the individual’s state over the previous week. Higher scores indicate greater well-being.

### Neuroticism measurements

Neuroticism was comprehensively assessed in PART wave III, as part of the Swedish-translated Schafer’s Five Factor Model (FFM) personality scale ^28-31^, which evaluates the ‘Big Five’ personality traits, i.e., extraversion, agreeableness, conscientiousness, openness, and neuroticism. Although the FFM scale was not incorporated in the previous PART waves, PART wave II included 14 neuroticism-related items from the Karolinska Scales of Personality (KSP). Specifically, the KSP inventory features items related to psychic anxiety, somatic anxiety, muscular tension, psychasthenia (lack of energy), inhibition of aggression (lack of assertiveness), guilt, suspicion, indirect aggression and irritability, all of which correlate positively with neuroticism in the Eysenck Personality Questionnaire-Revised (EPR-R) scale ^32^. FFM- and KSP-derived neuroticism scores were standardized into z-scores, and there was a significant positive correlation between them (Pearson’s r = 0.571, p < 0.001).

### Childhood adversities

Information on childhood adversities was obtained from the first wave of PART (PART wave I; 1998–2000) ^33^. These adversities, experienced before the age of 18, included severe family conflicts, significant financial hardships in the family, parental divorce, or the loss of a parent, and were coded categorically as either the absence of adversities or the presence of at least one. Recognizing the established association of childhood adversities with BMI ^34,35^, substance use ^36,37^, subjective well-being ^38,39^, and neuroticism ^40,41^, we accounted for them as covariates in our regression analyses (see below).

### Genotyping

A sub-cohort of the PART study provided DNA samples via self-administered saliva collection kits (Oragene DNA sample collection kit; DNA Genotek Inc., Stittsville, ON, Canada), as previously described ^42,43^. The rs1421085 SNP in *FTO* was genotyped using the TaqMan Universal Master Mix and a TaqMan SNP genotyping assay on an ABI 7900 HT instrument (Thermo Fisher Scientific, Waltham, MA, USA). This SNP was chosen based on its functional significance according to the RegulomeDB, the Brain eQTL Almanac (Braineac) and the GTEx database, as detailed in the Results section.

### Statistical analyses

Bivariate correlations between continuous variables were assessed using Pearson’s correlation coefficient. Linear regressions were used to examine the association of rs1421085 with neuroticism, well-being, BMI, and AUDIT scores in the PART cohort. The genotypic effects were modeled dominantly, with the T allele posited as the effect allele based on previous GWAS findings ^3^. Multicollinearity diagnostics were conducted using variance inflation factor (VIF) and tolerance metrics. Interaction terms were included in the regression models as appropriate. Cross-sectional and lagged longitudinal mediation analyses utilized Model 4 of the PROCESS macro for SPSS ^44^, estimating indirect effects via bias-corrected bootstrap confidence intervals from 5,000 bootstrap samples. Statistical analyses were performed using IBM SPSS Statistics v.27 (IBM Corp, Armonk, NY, USA), with alpha set at 0.05.

## Results

### PheWAS analysis reveals *FTO* gene associations across psychiatric traits

The phenome-wide association study (PheWAS) of the *FTO* gene considered 4,756 GWAS datasets encompassing a total of 3,302 phenotypic traits and yielded nominally significant associations with 791 traits across 27 domains (Fig. S1). Out of these, 242 traits across 17 domains met the criteria for multiple-testing corrected significance (Table S1). The most pronounced PheWAS signals for *FTO* resided within the metabolic domain and included body mass index (BMI), obesity status, waist-hip ratio, and various bioelectrical impedance measures (Fig. S1 and Table S1). Within the psychiatric domain, 26 traits survived multiple testing and were grouped into four categories (Table 1). More specifically, as shown in Table 1, the substance use category was the most prominent, highlighted by a leading association with alcohol consumption. The three other categories included traits related to chronotype/sleep, well-being, and neuroticism. There was also one trait related to a symptom of depression, i.e., recent poor appetite or overeating, which we attributed to the metabolic domain given its connection to caloric intake. In summary, the PheWAS findings underscore the diverse impact of the *FTO* gene, revealing its significant associations with a broad spectrum of psychiatric traits, most notably in substance use, chronotype and sleep, subjective well-being, and neuroticism, thus extending its known influence beyond metabolic traits.

**Table 1.** PheWAS of the *FTO* gene reveals associations with psychiatric traits related to substance use, chronotype/sleep, well-being, and neuroticism.

| Trait<sup>a</sup> | P-value<sup>a</sup> | N<sup>b</sup> | Category<sup>c</sup> |
| --- | --- | --- | --- |
| Average weekly beer plus cider intake | 3.12E-21 | 274556 | Substance Use |
| Chronotype | 7.52E-21 | 449732 | Chronotype/sleep |
| Morningness | 3.94E-18 | 345552 | Chronotype/sleep |
| Morning/evening person (chronotype) | 9.08E-18 | 345148 | Chronotype/sleep |
| Morning person (binary) | 5.11E-17 | 403195 | Chronotype/sleep |
| Sleep duration | 6.17E-14 | 384225 | Chronotype/sleep |
| Sleep duration | 6.97E-14 | 446118 | Chronotype/sleep |
| Sleep duration | 1.45E-13 | 384317 | Chronotype/sleep |
| Alcohol intake frequency | 5.15E-13 | 386082 | Substance Use |
| Ease of getting up in the morning | 2.78E-10 | 385949 | Chronotype/sleep |
| Getting up in morning | 4.15E-10 | 385494 | Chronotype/sleep |
| Snoring | 1.71E-09 | 359916 | Chronotype/sleep |
| Snoring | 2.69E-09 | 359498 | Chronotype/sleep |
| Depression - Recent poor appetite or overeating | 1.07E-08 | 126639 | Metabolic domain |
| Long sleep | 8.62E-08 | 339926 | Chronotype/sleep |
| Ever smoker | 2.61E-07 | 518633 | Substance Use |
| Drinks per day | 4.26E-07 | 537349 | Substance Use |
| Alcohol dependence | 5.15E-07 | 2322 | Substance Use |
| Happiness and subjective well-being - General happiness with own health | 5.85E-07 | 126477 | Well-being |
| Drinks per week | 9.08E-07 | 414343 | Substance Use |
| 10 mg response to amphetamine | 3.8017E-06 | 381 | Substance Use |
| Sleep duration (mean) | 8.7287E-06 | 85449 | Chronotype/sleep |
| Irritability | 9.2669E-06 | 369232 | Neuroticism |
| Past tobacco smoking | 2.1418E-05 | 355594 | Substance Use |
| Ever smoked | 3.5718E-05 | 385013 | Substance Use |
| Neuroticism | 3.958E-05 | 390278 | Neuroticism |

<sup>‡</sup> Only showing the traits from the psychiatric domain that passed the Bonferroni corrected P-value of 6.32e-5
<sup>a</sup> P-values are ranked from lowest to highest
<sup>b</sup> Total sample size in respective GWAS
<sup>c</sup> Traits were classified into four psychiatric categories, i.e., substance use, chronotype/sleep, well-being, and neuroticism. One depression-related symptom (i.e., recent poor appetite or overeating) was attributed to the metabolic domain given its relationship to caloric intake

### rs1421085: A putative regulatory variant within the *FTO* gene

Next, we aimed at replicating the associations of *FTO* with psychiatric traits by identifying and genotyping proxy SNP(s) with putative functionality. Among the eight SNPs associated with substance use phenotypes in GWAS studies (i.e., rs1421085, rs62033408, rs9937709, rs7188250, rs11642015, rs62048402, rs1558902, rs1477196) ^3-8^, rs1421085 had the highest possible functional score on RegulomeDB (probability score ≈ 1, ranking score: 1a; Table S2). Complementing this finding, the Brain eQTL Almanac (Braineac) provided supportive evidence for rs1421085 influencing *FTO* gene expression in neural tissues. Specifically, the T-allele, showed a nominal increase in *FTO* expression within the occipital cortex and cerebellum (Fig. S2), aligning with previous GWAS findings that identified the T allele as influential ^3^. The GTEx portal’s data also provided evidence for a regulatory role of rs1421085 in *FTO* expression in muscle, small intestine, pancreas, skin, and thyroid tissues (Fig. S3). It is also worth noting the significant correlation between rs1421085 and the other substance use-related SNPs, as established through linkage disequilibrium (LD) statistics. The LD measures generated by the LDpair Tool for the European population demonstrated a high degree of correlation with rs62033408 (D’: 1.0, R2: 0.918, p<0.0001), rs9937709 (D’: 0.950, R2: 0.874, p<0.0001), rs7188250 (D’: 0.937, R2: 0.840, p<0.0001), rs11642015, rs62048402, and rs1558902 (D’: 1.0, R2: 1.0, p<0.0001 for all). However, it was less correlated with rs1477196 (D’: 1.0, R2: 0.41, p<0.0001). Collectively, these data suggest that rs1421085 is an *FTO* variant with likely functional consequences and a suitable proxy in subsequent genotyping studies.

### Demographic and genotypic characteristics of the replication cohort

For our replication study, we leveraged data from the PART study which includes longitudinal measures on BMI, alcohol use, subjective well-being, and neuroticism (data on chronotype/sleep were not available). We genotyped rs1421085 in a cohort of 2,194 PART participants with females representing 60.3% of the total sample. The average age of genotyped participants was 56.6 years, with a standard deviation (SD) of 11.9 years, and a range from 31 to 77 years. The genotype distribution for rs1421085 was consistent with the Hardy-Weinberg equilibrium (χ^2^ = 0.0097, p = 0.92). Moreover, the observed genotype frequencies [TT: 810 individuals (36.9%), CT: 1,048 individuals (47.8%), CC: 336 individuals (15.3%)] aligned with the allele frequencies reported in the broader population by dbGaP (T allele frequency = 0.608, C allele frequency = 0.391, based on a sample size of 299,532 individuals; source: Alpha Allele Frequency release version: 20201027095038). Collectively, the genotypic data reinforce the representativeness of the replication cohort for investigating the genetic influences of the *FTO* variant rs1421085 on psychiatric traits.

### Validation of *FTO*’s associations with neuroticism and subjective well-being

In the replication cohort from the PART study, we started by exploring the association between the *FTO* gene and selected psychiatric-related traits, including alcohol use, neuroticism, and subjective well-being. We conducted linear regression analyses using the rs1421085 genotype under a dominant model of genetic effect, where the T allele was considered the effect allele according to previous GWAS findings ^3^. The analysis revealed that carriers of the T allele demonstrated significantly higher neuroticism scores; a finding that was consistent in both the unadjusted model (Table 2; model a, B: 0.12, p = 0.04) and the model accounting for age, sex, BMI, alcohol use, and childhood adversities (Table 2; model b, B: 0.16, p = 0.007). Upon examining subjective well-being, a significant inverse association with the rs1421085 genotype emerged in the model adjusted for age, sex, BMI, alcohol use, and childhood adversities, where T-allele carriers reported lower well-being levels (Table 3; model b, B: -0.90, p = 0.01). Notably, this association did not reach significance in the unadjusted analysis (Table 3; model a, B: -0.6, p = 0.1). In contrast, alcohol use, as measured by the AUDIT scales, did not show a significant association with rs1421085 in any model (Table S3). As a confirmatory analysis, we also examined the association between rs1421085 and BMI, and found it to be significant in both unadjusted and adjusted models, with T-allele carriers having lower BMI (Table S4). In summary, this validation phase extends the known association between *FTO* and BMI, to also include associations with higher levels of neuroticism and reduced subjective well-being, affirming the gene’s significant role in these psychiatric-related dimensions.

**Table 2.** Linear regression of neuroticism on the *FTO* rs1421085 genotype^‡^.

| N | B (95% CI)<sup>a</sup> | P | N | B (95% CI)<sup>b</sup> | P | N | B (95% CI)<sup>c</sup> | P |
| --- | --- | --- | --- | --- | --- | --- | --- | --- |
| 2,150 | 0.12 (0.002, 0.24) | 0.04 | 1,890 | 0.16 (0.04, 0.28) | 0.007 | 1,819 | 0.08 (-0.02, 0.18) | 0.11 |

B: Unstandardized beta coefficient, CI: Confidence Interval, P: p-value
<sup>‡</sup> T-allele carriers (CT or TT; coded 1) compared to CC homozygotes (CC; coded 0)
<sup>a</sup> Crude regression
<sup>b</sup> Adjusted for age, sex, BMI, AUDIT-10, childhood adversities
<sup>c</sup> Adjusted for age, sex, BMI, AUDIT-10, childhood adversities, well-being

**Table 3.** Linear regression of subjective well-being on the *FTO* rs1421085 genotype^‡^.

| N | B (95% CI)<sup>a</sup> | P | N | B (95% CI)<sup>b</sup> | P | N | B (95% CI)<sup>c</sup> | P |
| --- | --- | --- | --- | --- | --- | --- | --- | --- |
| 2,094 | -0.60 (-1.33, 0.12) | 0.10 | 1,847 | -0.90 (-1.64, -0.16) | 0.01 | 1,819 | -0.29 (-0.90, 0.30) | 0.33 |

B: Unstandardized beta coefficient, CI: Confidence Interval, P: p-value
<sup>‡</sup> T-allele carriers (CT or TT; coded 1) compared to CC homozygotes (CC; coded 0)
<sup>a</sup> Crude regression
<sup>b</sup> Adjusted for age, sex, BMI, AUDIT-10, childhood adversities
<sup>c</sup> Adjusted for age, sex, BMI, AUDIT-10, childhood adversities, neuroticism

### Exploring the indirect role of neuroticism in *FTO*’s association with well-being

Our replication efforts confirmed the connection between the *FTO* gene and both neuroticism and subjective well-being. As expected, there was a significant negative correlation between neuroticism and well-being within our cohort (Pearson’s r = -0.61, p < 0.001). To discern whether *FTO*’s associations with neuroticism and well-being were independent, we adjusted for one trait while analyzing the other. Interestingly, the significance of the rs1421085 *FTO* genotype on neuroticism disappeared when adjusting for well-being, and likewise, the *FTO* genotype’s significance on well-being disappeared when adjusting for neuroticism (Table 2; model c, B: 0.08, p = 0.11, and Table 3; model c, B: -0.29, p = 0.33; respectively). We further investigated the potential for neuroticism and well-being to either mediate or moderate the other’s effect in relation to the *FTO* genotype. Interaction terms were introduced in our regression models to assess moderation. These terms did not yield significant results, suggesting that neuroticism and well-being do not moderate the effect of the *FTO* genotype on each other (Tables S5 and S6). However, both cross-sectional and lagged longitudinal mediation analyses uncovered a significant indirect effect of rs1421085 on reduced well-being, which was mediated by higher neuroticism, in the models adjusted for age, sex, BMI, alcohol use, and childhood adversities (Table 4; cross-sectional, model b, Boot: -0.587, 95% CI [-1.040, -0.149]; longitudinal, model c, Boot: -0.433, 95% CI [-0.791, -0.069]). Although there was also a significant effect when examining the impact of rs1421085 on neuroticism through well-being in the cross-sectional adjusted mediation analysis, no such effect was observed in the lagged longitudinal analyses (Table S7).

**Table 4.** Indirect effect of the *FTO* rs1421085 genotype^‡^ on well-being (PART wave III) through neuroticism measured in PART wave III* or wave II**.

| N | Indirect Effect<br>(Boot 95% CI)<sup>a</sup> | Sig | N | Indirect Effect<br>(Boot 95% CI)<sup>b</sup> | Sig |
| --- | --- | --- | --- | --- | --- |
| 2,056 | -0.418 (-0.870, 0.033)<sup>*, a</sup> | No | 1,819 | -0.588 (-1.034, -0.143)<sup>*, b</sup> | Yes |
| 2,062 | -0.250 (-0.625, 0.133)<sup>**, a</sup> | No | 1,779 | -0.433 (-0.786, -0.079)<sup>**, c</sup> | Yes |

Boot CI: Bootstrap Confidence Interval, Sig: Statistical significance
<sup>‡</sup> T-allele carriers (CT or TT; coded 1) compared to CC homozygotes (CC; coded 0)
<sup>\*</sup> Cross-sectional mediation analysis
<sup>\*\*</sup> Lagged longitudinal mediation analysis
<sup>a</sup> Crude model
<sup>b</sup> Adjusted for age, sex, BMI (PART wave III), AUDIT-10 (PART wave III), childhood adversities
<sup>c</sup> Adjusted for age, sex, BMI (PART wave II and III), AUDIT-10 (PART wave II and III), childhood adversities

### Exploring the indirect role of BMI in *FTO*’s association with well-being

Next, since BMI has been causally linked to well-being ^45,46^ and since there was a modest but significant negative correlation between BMI and well-being in our replication cohort (Pearson’s r = -0.04, p < 0.001), we also examined the indirect effect of rs1421085 on well-being through BMI. Both cross-sectional and lagged longitudinal mediation analyses revealed a significant indirect effect of rs1421085 on enhanced well-being, which was mediated by lower BMI, in the models adjusted for age, sex, neuroticism, alcohol use, and childhood adversities (Table 5; cross-sectional, model b, Boot: 0.057, 95% CI [0.009, 0.127]; longitudinal, model c, Boot: 0.030, 95% CI [0.0004, 0.084]). There was no correlation between neuroticism and BMI in the replication cohort (Pearson’s r = -0.002, p = 0.93), and no mediating effect was observed for the impact of rs1421085 on neuroticism through BMI in cross-sectional and longitudinal analyses (Table S8). Taken together, the mediation results highlight a complex role of the *FTO* gene in affecting well-being, illustrating that while the rs1421085 T-allele may be associated with reduced well-being through elevated neuroticism, it simultaneously appears to be linked to improved well-being via a reduction in BMI.

**Table 5.** Indirect effect of the *FTO* rs1421085 genotype^‡^ on well-being (PART wave III) through BMI measured in PART wave III* or wave II* *.

| N | Indirect Effect<br>(Boot 95% CI) | Sig | N | Indirect Effect<br>(Boot 95% CI) | Sig |
| --- | --- | --- | --- | --- | --- |
| 2,075 | 0.028 (-0.004, 0.085)<sup>*, a</sup> | No | 1,819 | 0.058 (0.009, 0.133)<sup>*, b</sup> | Yes |
| 2,081 | 0.019 (-0.007, 0.067)<sup>**, a</sup> | No | 1,767 | 0.031 (0.0006, 0.083)<sup>**, c</sup> | Yes |

Boot CI: Bootstrap Confidence Interval, Sig: Statistical significance
<sup>‡</sup> T-allele carriers (CT or TT; coded 1) compared to CC homozygotes (CC; coded 0)
<sup>\*</sup> Cross-sectional mediation analysis
<sup>\*\*</sup> Lagged longitudinal mediation analysis
<sup>a</sup> Crude model
<sup>b</sup> Adjusted for age, sex, neuroticism (PART wave III), AUDIT-10 (PART wave III), childhood adversities
<sup>c</sup> Adjusted for age, sex, neuroticism (PART wave II and III), AUDIT-10 (PART wave II and III), childhood adversities

### Exploring the indirect role of neuroticism in *FTO*’s association with alcohol use

Direct associations between the *FTO* variant rs1421085 and alcohol use were not observed in the replication cohort (Table S2). However, given the association between rs1421085 and neuroticism (Table 2), and between neuroticism and alcohol use suggested by previous causal studies ^47^, we hypothesized that neuroticism could mediate an indirect association between rs1421085 and alcohol consumption. Both cross-sectional and lagged-longitudinal adjusted mediation analyses uncovered significant indirect associations between rs1421085 and problematic alcohol use (AUDIT-P), that operated via the influence of higher neuroticism (Table 6, AUDIT-P; cross-sectional, model b, Boot: 0.042, 95% CI [0.011, 0.080]; longitudinal, model c, Boot: 0.019, 95% CI [0.0002, 0.043]). Both crude and adjusted cross-sectional, but not longitudinal, analyses showed indirect associations between rs1421085 and AUDIT-10 via neuroticism (Table 6, AUDIT-10; cross-sectional, model a, Boot: 0.030, 95% CI [0.001, 0.071]; model b, Boot: 0.043, 95% CI [0.009, 0.088]). Interestingly, however, this mediating role of neuroticism did not extend to the AUDIT-C subscale, which does not capture problematic alcohol use (Table 6). We also examined indirect effects for rs1421085 on AUDIT scales through BMI but found no significances in the adjusted or lagged models (Table S9). In summary, while direct links between *FTO* and alcohol intake were not found in the replication cohort, our findings suggest again an indirect pathway, where higher neuroticism acts as a mediator between the *FTO* gene and problematic alcohol use.

**Table 6.**
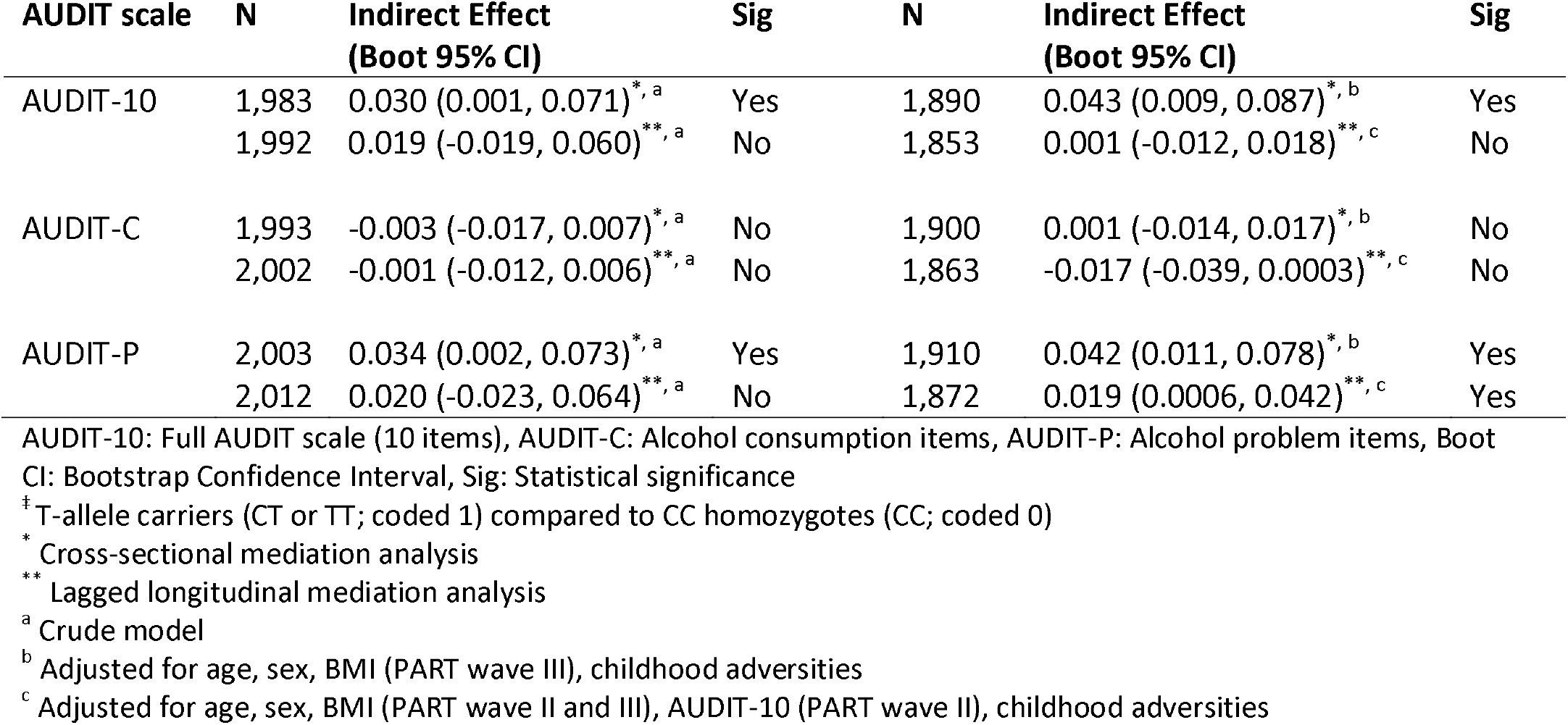
Indirect effect of the *FTO* rs1421085 genotype^‡^ on AUDIT scales (PART wave III) through neuroticism measured in PART wave III* or wave II* *.

## Discussion

Building upon the well-documented association between *FTO* gene variations and addiction phenotypes observed in the recent GWAS literature ^3-8^, including FTO’s involvement in dopaminergic signaling ^11^, our study probed for potential associations with a broader spectrum of psychiatric traits by means of a phenome-wide association study (PheWAS). The PheWAS for *FTO* uncovered a total of 242 traits across 17 domains that survived the multiple-correction testing criteria. The most pronounced signals resided within the metabolic domain, mirroring the findings from prior GWAS on obesity and related phenotypes ^1,2^. Within the psychiatric domain, the PheWAS revealed 26 significant psychiatric traits that belonged to four main categories, including (i) substance use, (ii) chronotype/sleep, (iii) well-being, and (iv) neuroticism. Connections between *FTO* and the first two categories have already been established by previous GWAS literature. Specifically, besides the already described *FTO* associations with substance use traits ^3-8^, previous GWAS studies have reported *FTO* associations with morningness and shorter sleep duration ^48-50^. However, the PheWAS findings on well-being and neuroticism are less well-established, although some preliminary evidence exists. For instance, a recent Mendelian Randomization (MR) study suggested that the *FTO* gene contributes to the genetic variation of BMI that is causally linked to well-being ^45^. In addition, a genetic study of a Korean traditional medicine system, which categorizes people into four constitutional types, reported an association between *FTO* and the (So-Eum) type that is characterized by high neuroticism ^51^.

To corroborate the PheWAS findings, we aimed at replicating the associations of *FTO* with psychiatric traits using the PART study which had longitudinal data on BMI, alcohol use, well-being, and neuroticism (but not chronotype/sleep). To this end, we first set out to identify a proxy SNP with putative functionality that could be genotyped in this cohort. Among the eight *FTO* SNPs previously associated with substance use phenotypes in GWAS studies ^3-8^, rs1421085 had the highest possible functional score on RegulomeDB suggesting its involvement in gene regulation and expression; a finding that was also supported by the Braineac and GTEx databases. Moreover, rs1421085 was the only variant that emerged as significant on the cross-ancestry level in three separate GWAS of alcohol use traits, including AUD ^3^, problematic alcohol use ^8^, and drinks per week ^6^. MR findings also suggested that rs1421085 is the SNP with the largest contribution to the genetic variation in BMI that affects well-being ^45^.

In our replication cohort, the significant association of rs1421085 with BMI was reaffirmed in both crude and adjusted linear regression models, supporting the established data on *FTO*’s influence on body mass ^1,2^. When examining psychiatric traits, rs1421085 exhibited significant associations with neuroticism and well-being after adjusting for age, sex, BMI, alcohol use, and childhood adversities. Specifically, carriers of the T allele, previously implicated in a heightened risk for AUD ^3^, showed higher levels of neuroticism and diminished well-being. Neuroticism constitutes a personality trait that is characterized by negative affectivity, including anxiety, anger, and emotional instability ^52^. Therefore, it is noteworthy that FTO’s molecular target, i.e., RNA methylation, has been implicated in regulating stress response mechanisms in the brain ^53^. Moreover, previous literature has consistently found neuroticism to be the personality trait that is most highly and negatively associated with different components of well-being, including happiness, life satisfaction, and quality of life ^54^. An inverse significant correlation between neuroticism and well-being was also present in our replication cohort. Previous GWAS of neuroticism have found significant negative genetic correlations with subjective well-being, and MR analyses have supported the presence of bidirectional associations between neuroticism and well-being ^55^. In the replication cohort, we investigated the potential for neuroticism and well-being to either mediate or moderate the other’s effect in relation to the *FTO* genotype. Through adjusted cross-sectional and longitudinal mediation analyses, we discerned a significant indirect effect of rs1421085 on well-being, with neuroticism acting as the mediator. Specifically, the T-allele of rs1421085 was implicated in decreased well-being through its influence on increased neuroticism. Next, considering the MR-suggested causal connection between BMI and well-being implicating rs1421085 ^45^, we extended our mediation inquiries to encompass BMI. In line with the MR study ^45^, our cross-sectional and lagged longitudinal mediation analyses uncovered a significant indirect influence of rs1421085 on subjective well-being, mediated by BMI. Specifically, the T-allele of rs1421085 was linked to enhanced well-being through its association with lower BMI. Taken together, our findings suggest that the T-allele’s effect on well-being is multifaceted, possibly involving a complex interplay between additional genetic predispositions and psychological traits.

Moreover, contrary to our expectations, our replication cohort did not demonstrate a direct link between rs1421085 and alcohol consumption. Building on the association between rs1421085 and neuroticism observed in our study, and between neuroticism and alcohol use observed in previous studies ^47^, we explored the concept of significant mediated effects in the absence of direct effects ^56^. Our adjusted cross-sectional and lagged longitudinal analyses, revealed that neuroticism significantly mediated the relationship between rs1421085 and the AUDIT-P subscale, which focuses on problematic alcohol use. Interestingly, this mediation was not observed with the AUDIT-C subscale that measures alcohol consumption levels without the problematic aspect. Given neuroticism’s role in predisposing individuals to various psychopathologies, it has been suggested that maladaptive substance use may be an attempt to mitigate the heightened anxiety, dysphoria, and emotional instability associated with this personality trait ^52^. In line with this assumption, recent MR studies have suggested a causal relationship between neuroticism and alcohol consumption ^47^. Past research on mediation has suggested that additional psychological constructs, like well-being ^27^ and coping strategies ^57^, can also indirectly influence the association between genetic predispositions and drinking behaviors. Taken together, these findings underscore the potential of genetic factors to precipitate problematic alcohol use by influencing psychological mechanisms of negative reinforcement, which are central to the development of alcohol use disorders ^58^.

In sum, our study is the first to establish a direct link between the *FTO* gene and neuroticism. Our data also indicate that *FTO* can indirectly influence well-being and problematic alcohol use through heightened neuroticism. These findings may have significant public health implications. Specifically, they position the FTO enzyme as a promising target for therapeutic interventions and highlight the potential of focusing on neuroticism—a trait putatively susceptible to treatment ^59^—as a strategy to alleviate psychopathology. To confirm and expand our understanding of these findings, further longitudinal and translational studies are needed.

## Data Availability

All data produced in the present study are available upon reasonable request to the authors

## Acknowledgements

This work was supported by the Royal Physiographic Society in Lund (42797, 2022; P.A.M.), the Åke Wiberg Foundation (M21-0092, 2021 and M22-0059, 2022; P.A.M.), the Magnus Bergvall Foundation (2021-04173, 2021 and 2022-038, 2022; P.A.M.), the Sigurd and Elsa Golje Memorial Foundation (LA2021-0124, 2021 and LA2022-0139; P.A.M.), the Swedish Brain Foundation (FO2023-0167; P.A.M. and 2023-0335; C.L.), the Karolinska Institutet Research Grants (2022-01667, 2022; P.A.M.), the regional agreement on medical training and clinical research (ALF) between Stockholm County Council and Karolinska Institutet Stockholm County Council (RS2021-0855, SLL20190589; C.L.), and the Swedish Research Council (2023-02253; P.A.M., and 2022-1188; C.L.). The funding sources were not involved in the study design or the decision to submit the results for publication.

## Author contributions

W.C.: Investigation; methodology; writing—review and editing. Y.F.: Methodology; project administration; resources; writing—review and editing. C.L.: Funding acquisition; methodology, resources; supervision; writing—review and editing. P.A.M.: Conceptualization; data curation; formal analysis; funding acquisition; investigation; methodology; project administration; resources; supervision; validation; visualization; writing—original draft. All authors have read and agreed to the published version of the manuscript.

## Data availability statement

The data are not publicly available due to ethical and privacy restrictions. Data are available on reasonable request from the corresponding author.

