## Supplemental figures and tables for "Examining the association between the *FTO* gene and neuroticism reveals indirect effects on subjective well-being and problematic alcohol use"

Fig. S1

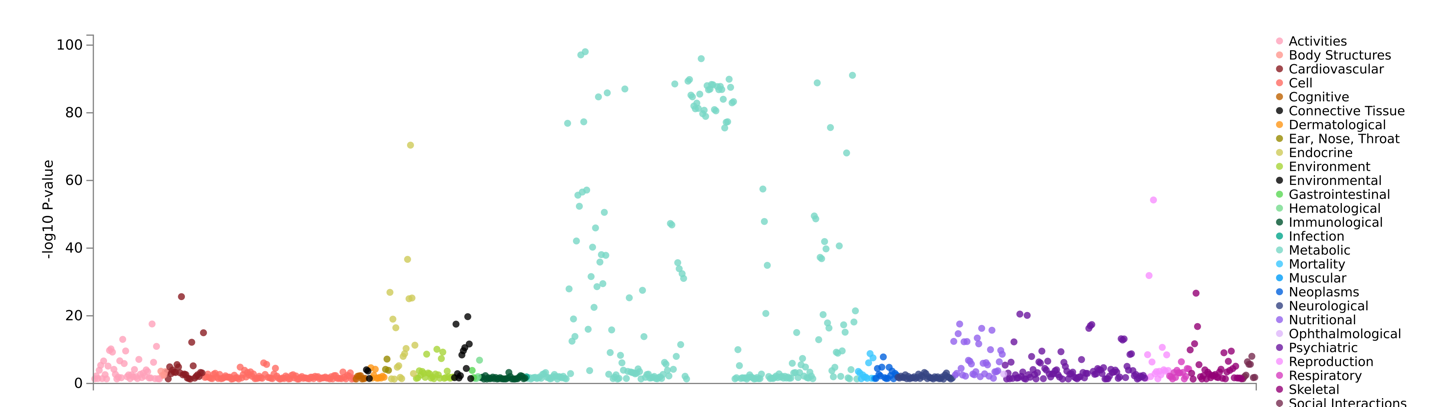

**Figure S1. PheWAS plot for the *FTO* gene**

The PheWAS considered 4,756 GWASs and found 791 significant traits (P<0.05) categorized in different domains (e.g., cardiovascular, dermatological, immunological, etc.). In the psychiatric domain, a total of 26 traits passed the multiple-corrected significance (Bonferroni corrected P-value: 6.32e-5; Table 1).

Fig. S2

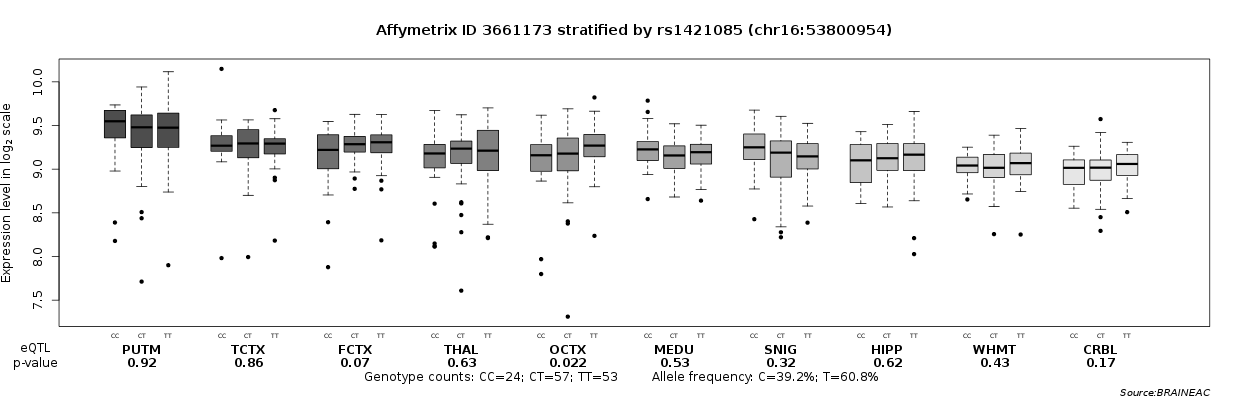

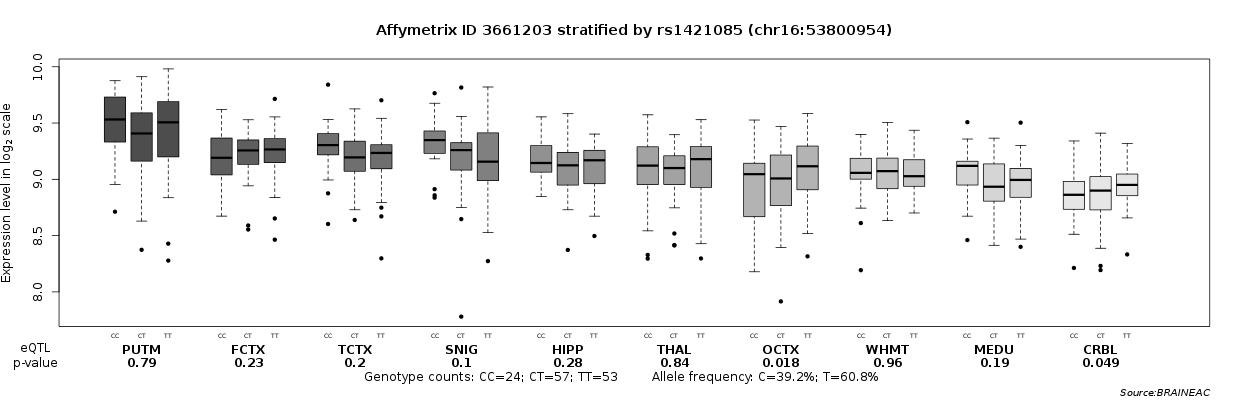

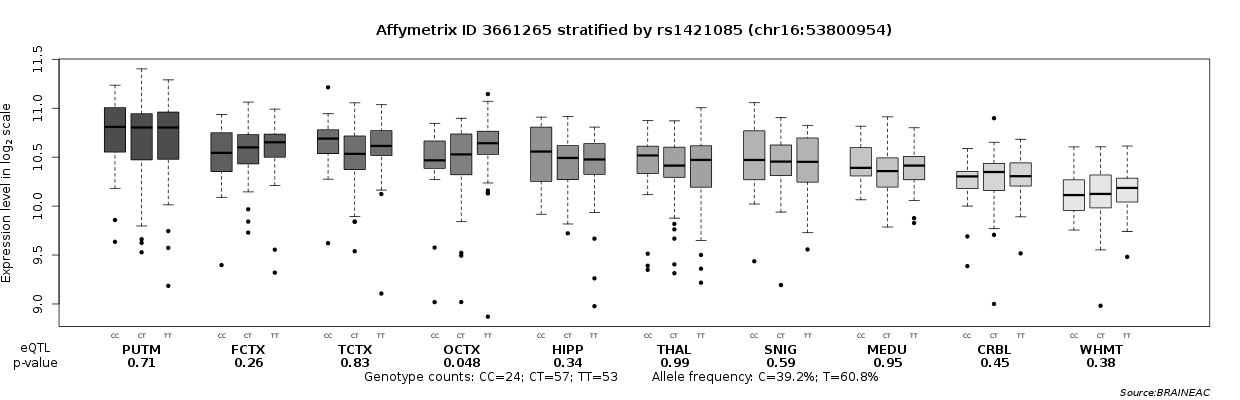

**Figure S2. Brain-region eQTL analyses for *FTO*’s rs1421085**

The UK Brain Expression Consortium (UKBEC) provides the Brain eQTL Almanac (Braineac; http://www.braineac.org), an online resource for exploring how genetic variation affects gene expression in 10 different regions of the human brain. Three out of 14 exon-specific probe sets provided suggestive evidence for a regulatory role of rs1421085 on *FTO* gene expression. Specifically, there were nominal eQTL significances for the occipital cortex (OCTX; p=0.022 for Affymetrix probe ID 3661173, p=0.018 for probe ID 3661203, and p=0.048 for probe ID 3661265) and the cerebellum (CRBL; p=0.049 for probe ID 3661203), indicative of the T-allele increasing *FTO* expression.

Fig. S3

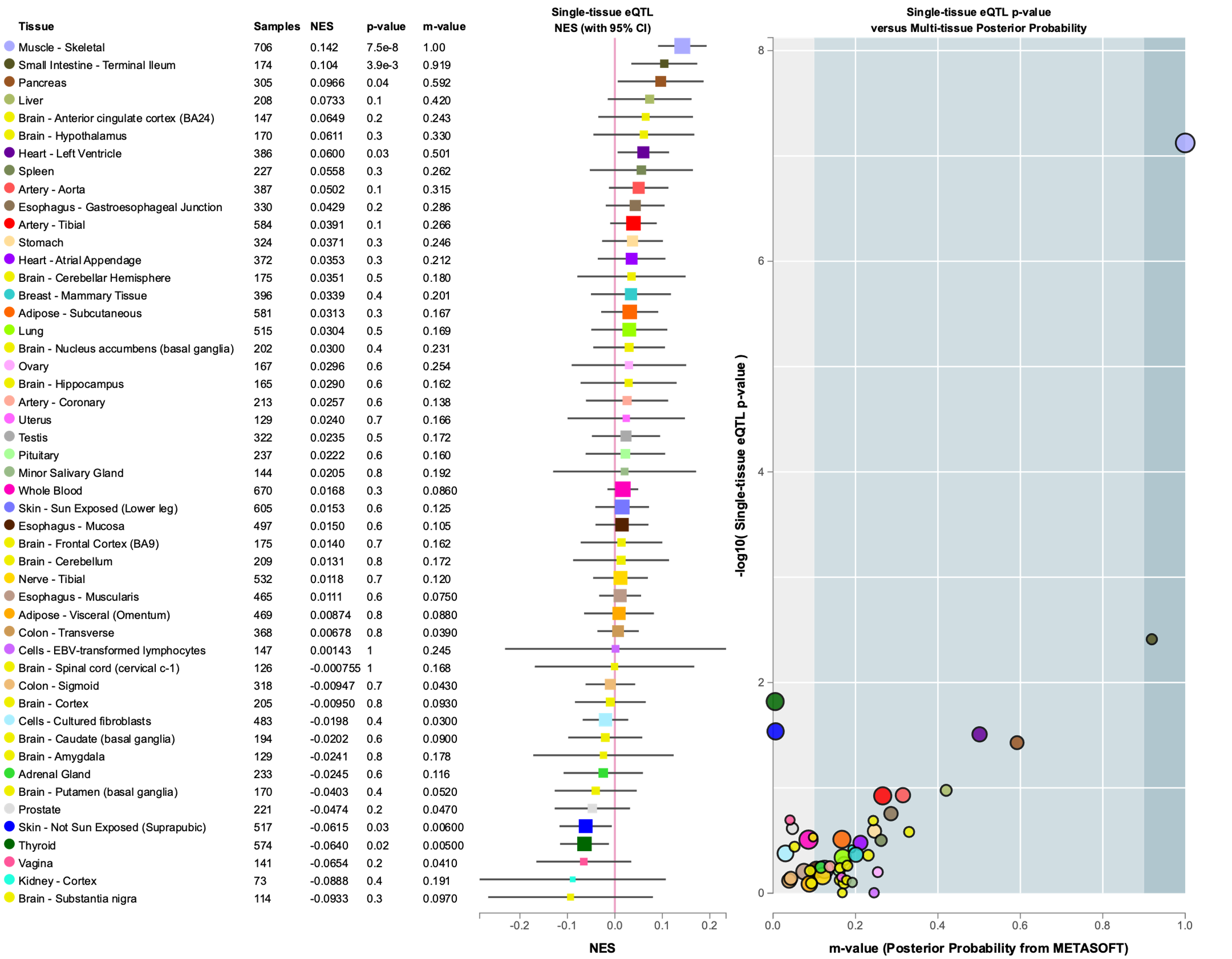

**Figure S3. Multi-tissue eQTL analyses for *FTO*’s rs1421085**

A multi-tissue eQTL plot was generated using the GTEx Portal (https://gtexportal.org/home/). The strongest evidence for a regulatory role of rs1421085 on *FTO* expression was in muscle (p=7.5e-8), and with suggestive evidence in small intestine (p=3.9e-3), pancreas (p=0.04), skin (p=0.03), and thyroid (p=0.02).

**Table S1. PheWAS for the *FTO* gene uncovered 242 traits across 17 domains that met the criteria for multiple-testing corrected significance**

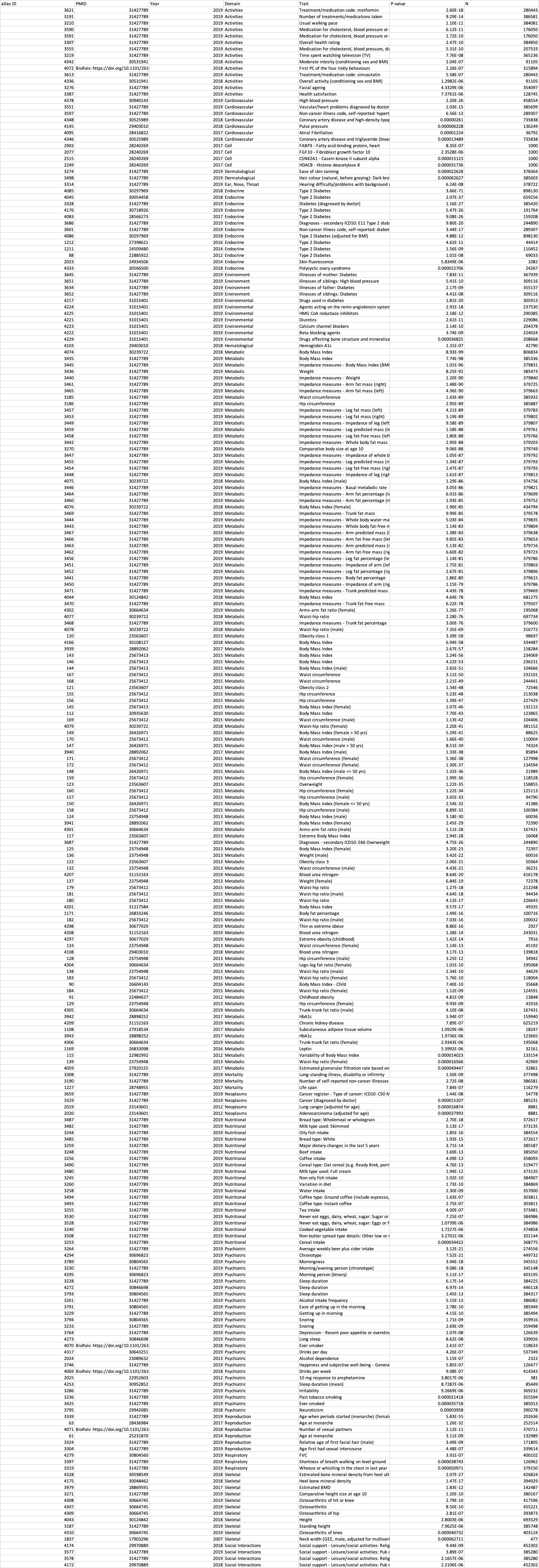

**Table S2. RegulomeDB results for the eight *FTO* SNPs associated with substance use phenotypes in GWAS studies**

| **FTO SNPs** | **Probability score*** | **Ranking**** |
| --- | --- | --- |
| rs1421085 | 0.97 | 1a |
| rs11642015 | 0.55436 | 1f |
| rs62048402 | 0.66703 | 1f |
| rs1558902 | 0.22271 | 1f |
| rs1477196 | 0.8313 | 1f |
| rs9937709 | 0.05 | 1f |
| rs62033408 | 0.66703 | 1f |
| rs7188250 | 0.55324 | 1f |

*The RegulomeDB probability score is ranging from 0 to 1, with 1 being most likely to be a regulatory variant. The probabilistic score is calculated from a random forest model, TURF, trained with allele-specific transcription factor (TF) binding single nucleotide variants (SNVs).

**The scoring scheme refers to the following supporting evidence for that particular location or variant id. In general, if more supporting data is available, the higher is its likelihood of being functional and hence receiving a higher score (with 1 being higher and 7 being lower score).

Score 1a: Supporting data includes eQTL/caQTL + TF binding + matched TF motif + matched Footprint + chromatin accessibility peak.

Score 1f data includes eQTL/caQTL + TF binding / chromatin accessibility peak.

For complete scoring of data see https://regulomedb.org/regulome-help/

**Table S3. Linear regression of AUDIT scales on the FTO rs1421085 genotype^‡^ in the PART cohort**

| **AUDIT scale** | **N** | **B (95% CI)^a^** | **P** | **N** | **B (95% CI)^b^** | **P** |
| --- | --- | --- | --- | --- | --- | --- |
| AUDIT-10 | 2,023 | -0.09 (-0.43, 0.25) | 0.59 | 1,819 | -0.03 (-0.37, 0.31) | 0.85 |
| AUDIT-C | 2,033 | -0.08 (-0.29, 0.12) | 0.43 | 1,828 | -0.07 (-0.29, 0.14) | 0.50 |
| AUDIT-P | 2,045 | -0.07 (-0.26, 0.12) | 0.47 | 1,834 | 0.03 (-0.14, 0.21) | 0.68 |

AUDIT-10: Full AUDIT scale (10 items), AUDIT-C: Alcohol consumption items, AUDIT-P: Alcohol problem items, B: Unstandardized beta coefficient, CI: Confidence Interval, P: p-value

^‡^ T-allele carriers (CT or TT; coded 1) compared to CC homozygotes (CC; coded 0)

^a^ Crude regression

^b^ Adjusted for age, sex, BMI, neuroticism, well-being, childhood adversities

**Table S4. Linear regression of BMI on the FTO rs1421085 genotype^‡^ in the PART cohort**

| **N** | **B (95% CI)^a^** | **P** | **N** | **B (95% CI)^b^** | **P** |
| --- | --- | --- | --- | --- | --- |
| 2,171 | -0.47 (-0.93, -0.01) | 0.04 | 1,819 | -0.61 (-1.10, -0.12) | 0.01 |

B: Unstandardized beta coefficient, CI: Confidence Interval, P: p-value

^‡^ T-allele carriers (CT or TT; coded 1) compared to CC homozygotes (CC; coded 0)

^a^ Crude regression

^b^ Adjusted for age, sex, AUDIT-10, neuroticism, well-being, childhood adversities

**Table S5. Linear regression of neuroticism on the interaction between the FTO rs1421085 genotype and subjective well-being in the PART cohort**

| **N** | **B (95% CI)^a^** | **P** |
| --- | --- | --- |
| 1,819 | 0.001 (-0.01, 0.01) | 0.91 |

B: Unstandardized beta coefficient, CI: Confidence Interval, P: p-value

^a^ Also included in the model: age, sex, BMI, AUDIT-10, well-being, rs1421085, childhood adversities

**Table S6. Linear regression of subjective well-being on the interaction between the FTO rs1421085 genotype and neuroticism in the PART cohort**

| **N** | **B (95% CI)^a^** | **P** |
| --- | --- | --- |
| 1,819 | -0.04 (-0.66, 0.57) | 0.88 |

B: Unstandardized beta coefficient, CI: Confidence Interval, P: p-value

^a^ Also included in the model: age, sex, BMI, AUDIT-10, neuroticism, rs1421085, childhood adversities

**Table S7. Indirect effect of the FTO rs1421085 genotype^‡^ on neuroticism (PART wave III) through well-being measured in PART wave III^*^ or wave II^**^**

| **N** | **Indirect Effect**  **(Boot 95% CI)^a^** | **Sig** | **N** | **Indirect Effect**  **(Boot 95% CI)^b^** | **Sig** |
| --- | --- | --- | --- | --- | --- |
| 2,056 | 0.060 (-0.011, 0.133)^*, a^ | No | 1,819 | 0.087 (0.015, 0.159)^*, b^ | Yes |
| 2,125 | 0.030 (-0.032, 0.091)^**, a^ | No | 1,827 | 0.048 (-0.011, 0.108)^**, c^ | No |

Boot CI: Bootstrap Confidence Interval, Sig: Statistical significance

^‡^ T-allele carriers (CT or TT; coded 1) compared to CC homozygotes (CC; coded 0)

^*^ Cross-sectional mediation analysis

^**^ Lagged longitudinal mediation analysis

^a^ Crude model

^b^ Adjusted for age, sex, BMI (PART wave III), AUDIT-10 (PART wave III), childhood adversities

^c^ Adjusted for age, sex, BMI (PART wave II and III), AUDIT-10 (PART wave II and III), childhood adversities

**Table S8. Indirect effect of the FTO rs1421085 genotype^‡^ on neuroticism (PART wave III) through BMI measured in PART wave III^*^ or wave II^**^**

| **N** | **Indirect Effect**  **(Boot 95% CI)** | **Sig** | **N** | **Indirect Effect**  **(Boot 95% CI)** | **Sig** |
| --- | --- | --- | --- | --- | --- |
| 2,123 | -0.0003 (-0.007, 0.005)^*, a^ | No | 1,819 | 0.004 (-0.001, 0.011)^*, b^ | No |
| 2,135 | -0.0005 (-0.005, 0.004)^**, a^ | No | 1,775 | 0.001 (-0.002, 0.008)^**, c^ | No |

Boot CI: Bootstrap Confidence Interval, Sig: Statistical significance

^‡^ T-allele carriers (CT or TT; coded 1) compared to CC homozygotes (CC; coded 0)

^*^ Cross-sectional mediation analysis

^**^ Lagged longitudinal mediation analysis

^a^ Crude model

^b^ Adjusted for age, sex, well-being (PART wave III), AUDIT-10 (PART wave III), childhood adversities

^c^ Adjusted for age, sex, well-being (PART II and III), AUDIT-10 (PART II and III), childhood adversities

**Table S9. Indirect effect of the FTO rs1421085 genotype^‡^ on AUDIT scales (PART wave III) through BMI measured in PART wave III^*^ or wave II^**^**

| **AUDIT scale** | **N** | **Indirect Effect**  **(Boot 95% CI)** | **Sig** | **N** | **Indirect Effect**  **(Boot 95% CI)** | **Sig** |
| --- | --- | --- | --- | --- | --- | --- |
| AUDIT-10 | 2,005 | -0.017 (-0.046, 0.0002)^*, a^ | No | 1,890 | -0.0004 (-0.019, 0.016)^*, b^ | No |
|  | 2,011 | -0.011 (-0.039, 0.004)^**, a^ | No | 1,832 | -0.001 (-0.014, 0.008)^**, c^ | No |
| AUDIT-C | 2,015 | -0.017 (-0.040, -0.001)^*, a^ | Yes | 1,900 | -0.007 (-0.023, 0.004)^*, b^ | No |
|  | 2,021 | -0.008 (-0.026, 0.002)^**, a^ | No | 1,842 | -0.002 (-0.013, 0.004)^**, c^ | No |
| AUDIT-P | 2,027 | 0.0008 (-0.009, 0.011)^*, a^ | No | 1,910 | 0.007 (-0.0006, 0.022)^*, b^ | No |
|  | 2,032 | -0.002 (-0.014, 0.007)^**, a^ | No | 1,849 | 0.001 (-0.004, 0.009)^**, c^ | No |

AUDIT-10: Full AUDIT scale (10 items), AUDIT-C: Alcohol consumption items, AUDIT-P: Alcohol problem items, Boot CI: Bootstrap Confidence Interval, Sig: Statistical significance

^‡^ T-allele carriers (CT or TT; coded 1) compared to CC homozygotes (CC; coded 0)

^*^ Cross-sectional mediation analysis

^**^ Lagged longitudinal mediation analysis

^a^ Crude model

^b^ Adjusted for age, sex, neuroticism (PART wave III), childhood adversities

^c^ Adjusted for age, sex, neuroticism (PART wave II and III), AUDIT-10 (PART wave II), childhood adversities
